# Data-driven decision support for individualised cardiovascular resuscitation in sepsis: a scoping review and primer for clinicians

**DOI:** 10.1101/2023.08.26.23294666

**Authors:** Finneas JR Catling, Myura Nagendran, Paul Festor, Zuzanna Bien, Steve Harris, A Aldo Faisal, Anthony C Gordon, Matthieu Komorowski

## Abstract

**Background:** We conducted a scoping review of machine learning systems that inform individualised cardiovascular resuscitation of adults in hospital with sepsis. Our study reviews the resuscitation tasks that the systems aim to assist with, system robustness and potential to improve patient care, and progress towards deployment in clinical practice. We assume no expertise in machine learning from the reader and introduce technical concepts where relevant.

**Methods:** This study followed the *Preferred Reporting Items for Systematic reviews and Meta-Analyses extension for Scoping Reviews* guidance. MEDLINE, EMBASE, Scopus, ClinicalTrials.gov, arXiv, bioRxiv and medRxiv were systematically searched up to September 2021. We present a narrative synthesis of the included studies, which also aims to equip clinicians with an understanding of the foundational machine learning concepts necessary to interpret them.

**Results:** 73 studies were included with 80% published after 2018. Supervised learning systems were often used to predict septic shock onset. Reinforcement learning systems were increasingly popular in the last five years, and were used to guide specific dosing of fluids and vasopressors. A minority of studies proposed systems containing biological models augmented with machine learning. Sepsis and septic shock were heterogeneously defined and 63% of studies derived their systems using a single dataset. Most studies performed only retrospective internal validation, with no further steps taken towards translating their proposed systems into clinical practice.

**Conclusions:** Machine learning systems can theoretically match, or even exceed, human performance when predicting patient outcomes and choosing the most suitable cardiovascular treatment strategy in sepsis. However, with some notable exceptions, the vast majority of systems to date exist only as proof of concept, with significant barriers to translation.

## Introduction

This study reviews data-driven systems used to inform individualised cardiovascular resuscitation of adult patients in hospital with sepsis [1]. The available systems performed varied tasks and were evaluated in differing patient cohorts and at different levels of fidelity. They were therefore not suitable for direct comparison. We thus performed a scoping review, seeking to address the following questions:

1. Which tasks within sepsis resuscitation did the systems aim to assist with, and what approaches (algorithms, patient cohorts and methods of validation) were used to produce them?
2. What were the advantages and disadvantages of the different systems in terms of their robustness (i.e. their integrity under different operating conditions [2]) and potential to improve patient care?
3. How far had the systems progressed towards deployment in clinical practice? Where systems had been deployed, what was their impact?

Machine learning systems are often highly technical, making their inner workings opaque to many healthcare professionals who are increasingly expected to use them. We aim to provide an accessible introduction to machine learning concepts necessary to interpret the output of these systems and avoid their pitfalls.

## Methods

This study was conducted in accordance with the Preferred Reporting Items for Systematic reviews and Meta-Analyses extension for Scoping Reviews (PRISMA-ScR) [3]. Our study protocol was prospectively registered with the Open Science Framework (see https://osf.io/c6ynq/) on 14 October 2021.

### Eligibility criteria

Studies were included that evaluated a data-driven system for individualised cardiovascular resuscitation in adult (18 years of age, or older) inpatients with sepsis. Eligible systems had to satisfy all three of the following criteria:

1. They recommended cardiovascular treatment (defined as intravenous fluids, vasopressors or inotropes), or predicted response to cardiovascular treatment with the intent of informing future cardiovascular treatment.
2. They contained components learnt directly from data, excluding clinical guidelines and ‘expert systems’ which contain only human-authored rules based on clinical experience or a review of relevant literature.
3. They made recommendations or predictions that changed depending on the values of one or more routinely-measured clinical variables.

Additional inclusion criteria included:

- Primary interventional or observational research, or service evaluation. Quantitative and qualitative approaches were both eligible.
- Manuscripts written in English or French.
- Data-driven systems for realtime early diagnosis of septic shock (defined as predicting development of septic shock within, at most, the next 48 hours). This was because many investigators treat a prescription of vasopressor treatment in sepsis as synonymous with a diagnosis of septic shock, resulting in systems which essentially predict vasopressor prescriptions in sepsis.
- Systems could be included where they used a simple human-authored rule to link a data-driven prediction of an outcome to a treatment recommendation, as long as knowledge of the predicted outcome mandated or strongly encouraged the recommended cardiovascular treatment.

Definitions of sepsis and septic shock have evolved over time, and have been adapted for use in different healthcare systems. We therefore included studies that use the Sepsis-1 [4], Sepsis-2 [5] or Sepsis-3 [1] definitions, as well as reasonable precursors or adaptations of these. Exclusion criteria included:

- Review articles, commentaries, editorials, case reports, conference abstracts without an associated conference paper, unmodified replications of previous studies and incomplete or unpublished clinical trials.
- Evaluations exclusively in paediatric populations, animals or synthetic data.
- Evaluations in general populations of adult inpatients, except where a subgroup analysis of exclusively the patients with sepsis was available.
- Studies that report associations between risk factors and outcomes without specifying how these should be used to inform treatment.
- Systems that use non-routinely-collected data, including transcriptomics or cytokine profiles.

### Literature search

MEDLINE, EMBASE, Scopus, ClinicalTrials.gov, arXiv, bioRxiv and medRxiv were searched from database inception to September 20th, 2021. The search strategies were drafted and refined in consultation with the Imperial College London library team, and in accordance with the Peer Review of Electronic Search Strategies (PRESS) Guideline [6]. Studies only in animals were excluded using the technique recommended in [7], with an adaptation to also exclude non-adults [8]. Our final search strategy is shown in Appendix A (Additional File 1). The retrieved studies were supplemented by scanning the reference lists of relevant review articles.

Article screening, decisions on inclusion and data extraction were performed collaboratively by five reviewers (FJRC, MN, ZB, PF and MK) using the Covidence systematic review software (Veritas Health Innovation, Melbourne, Australia), versions 2673 and above. Duplicate articles were removed using Covidence’s automated duplicate detection function. Each record was reviewed by at least two reviewers with consensus opinion from all co-authors sought on any inter-reviewer discrepancies. Single reviewers extracted data items from each article, flagging contentious areas for further scrutiny by two reviewers and consensus decision. The full screening and extraction process was preceded by a joint calibration exercise using 69 articles.

A data collection form (see Appendix B, Additional File 1) was produced prior to search execution. This included items on article characteristics (title, year, funding source), algorithms used, datasets (numbers of patients and hospitals, data resolution) and progress towards translation.

### Data synthesis

We grouped the included studies according to the main algorithmic approach used (supervised learning, unsupervised learning, reinforcement learning or biological modelling, as defined in the Results). We then conducted a narrative synthesis within each category, describing the general features of each approach, summarising common methodologies, and highlighting notable contributions from individual studies.

## Results

This section begins by summarising our main findings, then presents an introduction to the four major approaches used to build data-driven sepsis-resuscitation systems. This later material is intended to be accessible to a non-technical audience, and highlights the clinical relevance of individual studies.

We included 73 studies which evaluate systems for individualised resuscitation in sepsis. Study identification and screening is summarised in Figure 1, and characteristics of the included studies are presented in Table 1. All extracted study data are available in Additional File 2. 55 studies (75%) used supervised learning, typically for realtime septic shock prediction or in combination with reinforcement learning. 26 studies (36%) used reinforcement learning, all of which recommended treatment strategies using IV fluids and vasopressors. 8 studies (11%) used unsupervised learning and 4 (6%) used biological models augmented with machine learning. Most of the studies (58, 80%) were published in or after 2018, with only 3 (4%) studies published before 2010. The temporal trends in machine learning approach used are shown in Figure 2, which highlights the increasing popularity of reinforcement learning in recent years.

**Figure 1.**
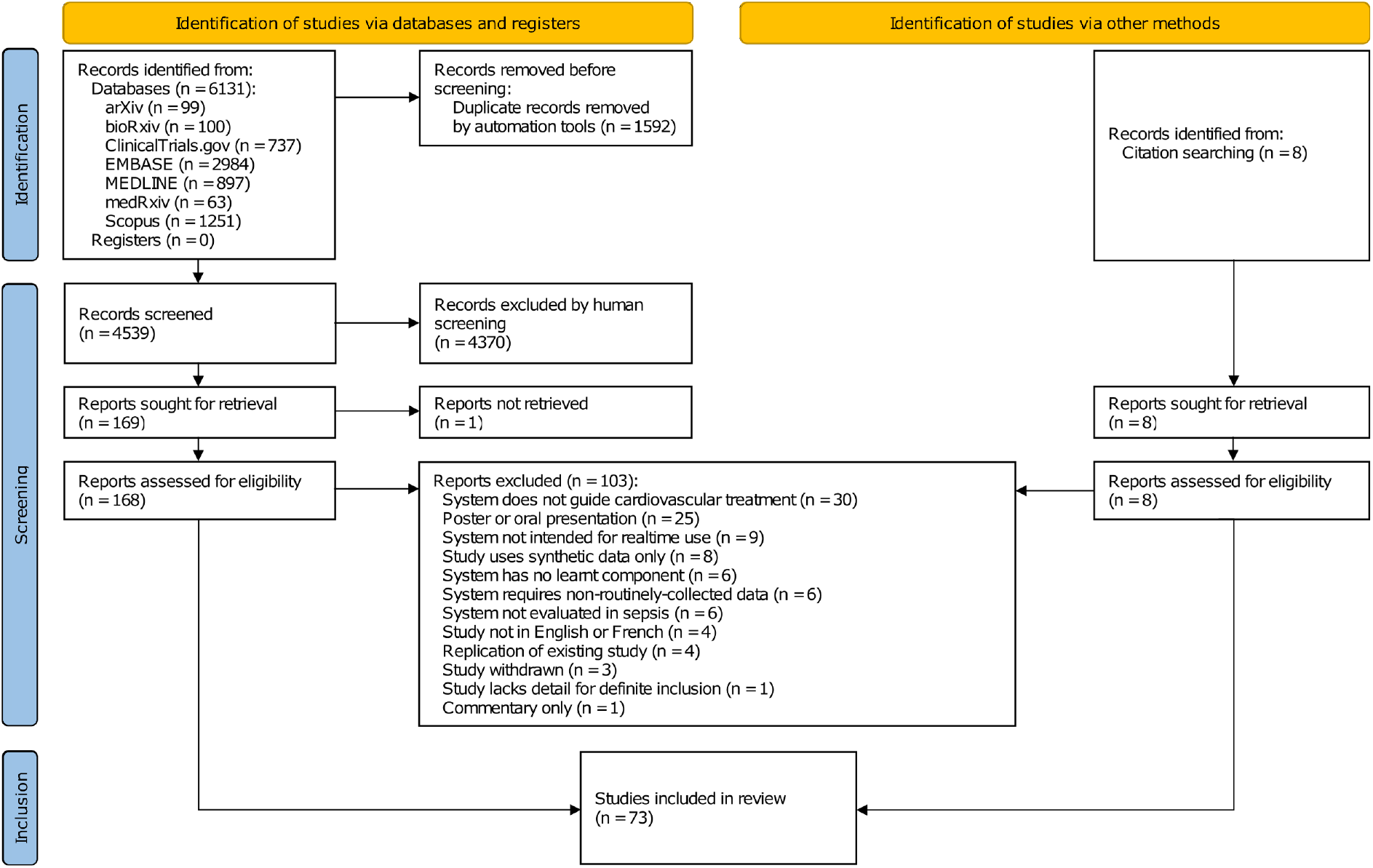
PRISMA flow diagram showing identification and screening of studies.

**Table 1.**
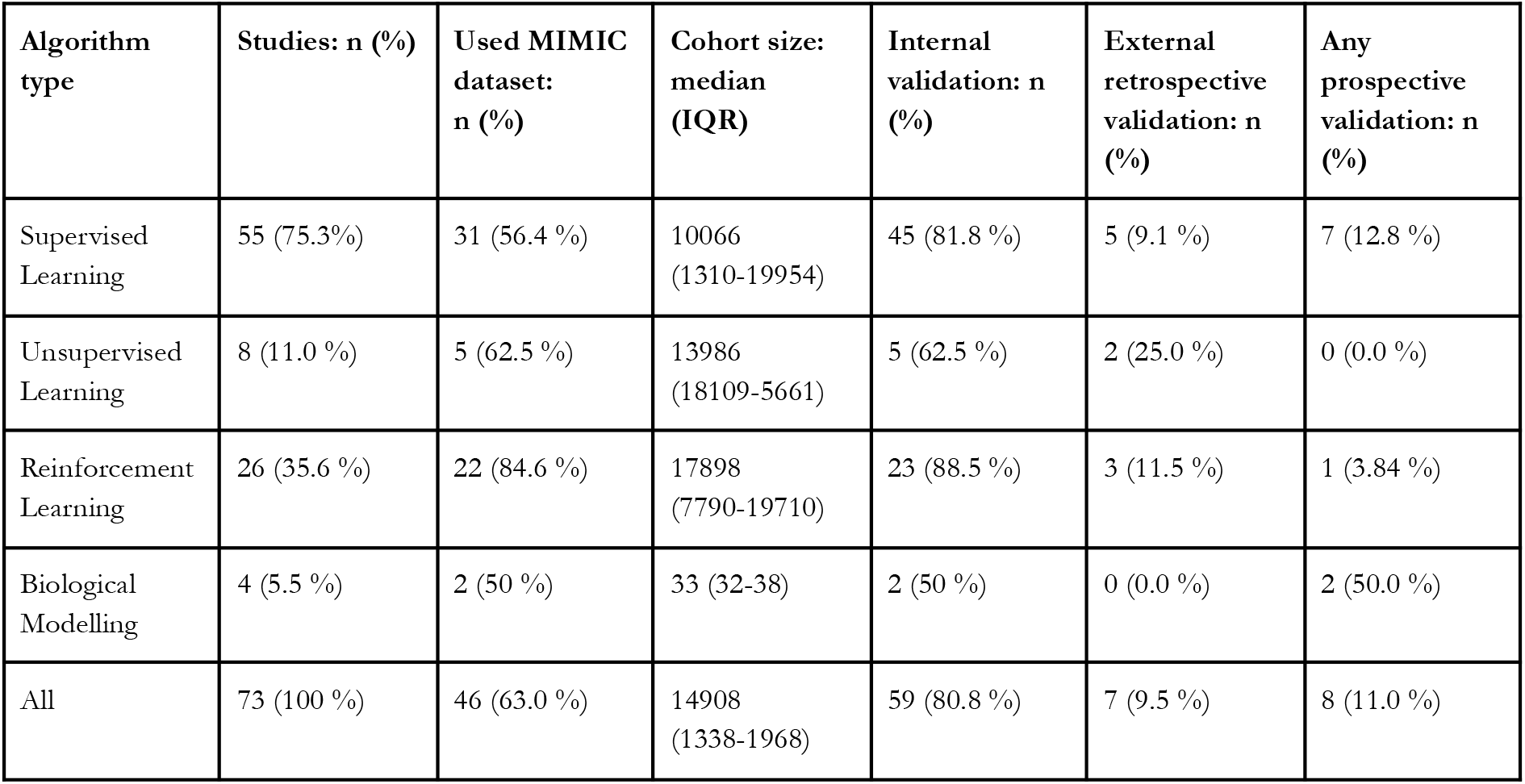
Characteristics of the included studies, stratified by algorithm type. Individual columns may sum to greater than the total in the final row as a single study may include more than one algorithm type.

| Algorithm type | Studies: n (%) | Used MIMIC dataset: n (%) | Cohort size: median (IQR) | Internal validation: n (%) | External retrospective validation: n (%) | Any prospective validation: n (%) |
| --- | --- | --- | --- | --- | --- | --- |
| Supervised Learning | 55 (75.3%) | 31 (56.4 %) | 10066 (1310-19954) | 45 (81.8 %) | 5 (9.1 %) | 7 (12.8 %) |
| Unsupervised Learning | 8 (11.0 %) | 5 (62.5 %) | 13986 (18109-5661) | 5 (62.5 %) | 2 (25.0 %) | 0 (0.0 %) |
| Reinforcement Learning | 26 (35.6 %) | 22 (84.6 %) | 17898 (7790-19710) | 23 (88.5 %) | 3 (11.5 %) | 1 (3.84 %) |
| Biological Modelling | 4 (5.5 %) | 2 (50 %) | 33 (32-38) | 2 (50 %) | 0 (0.0 %) | 2 (50.0 %) |
| All | 73 (100 %) | 46 (63.0 %) | 14908 (1338-1968) | 59 (80.8 %) | 7 (9.5 %) | 8 (11.0 %) |

**Figure 2.**
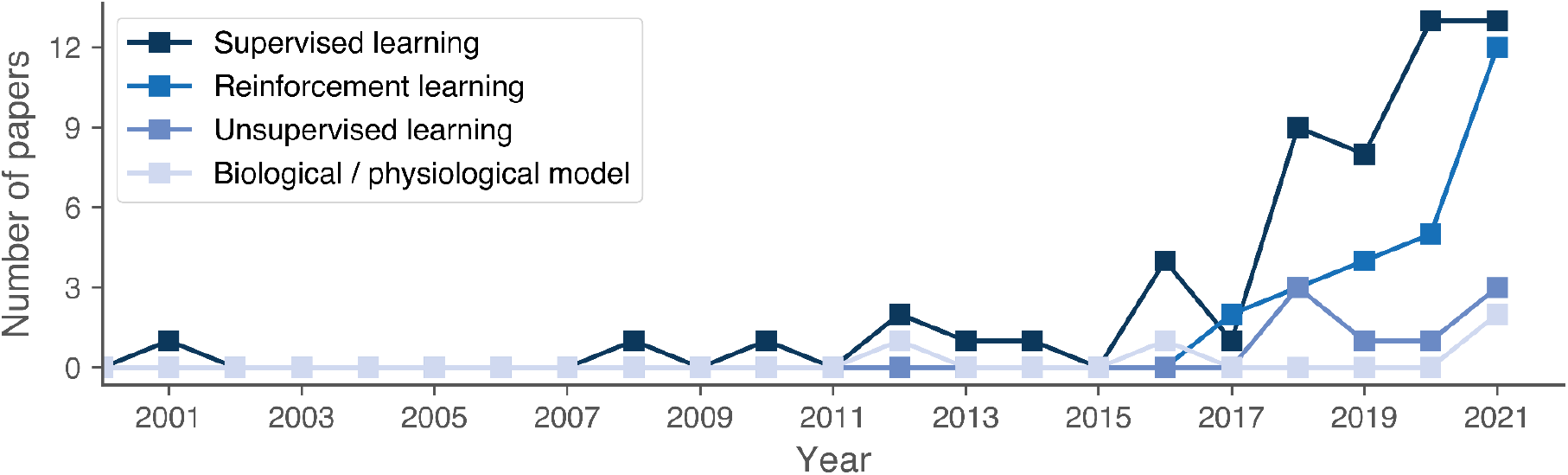
Trends in machine learning approaches used, over time.

Most studies performed only retrospective internal validation. As summarised in Figure 3 (using the format proposed in [9]), significant further steps would thus be required before most systems could be deployed in clinical practice. Three of the four [10–13] systems that had progressed to interventional trials either had a minimal learnt component [10] or were constrained by physiological principles [11,12]. The majority of studies received academic funding, although private funding was increasingly common in recent studies, indicating a commercial interest in data-driven sepsis resuscitation (see Supplementary Figure 1, Additional File 1). Sepsis was heterogeneously defined across the studies (see Supplementary Figure 2, Additional file 1): 34 (47%) adopted a definition based on the ‘sepsis 3’ criteria [1], whilst 14 (19%) used clinical codes and 13 (18%) used evidence of infection plus a systemic inflammatory response [5]. Of the studies that defined septic shock, just 17 (52%) used a definition requiring initial fluid resuscitation, and only 12 (36%) used a definition requiring elevated lactate.

**Figure 3.**
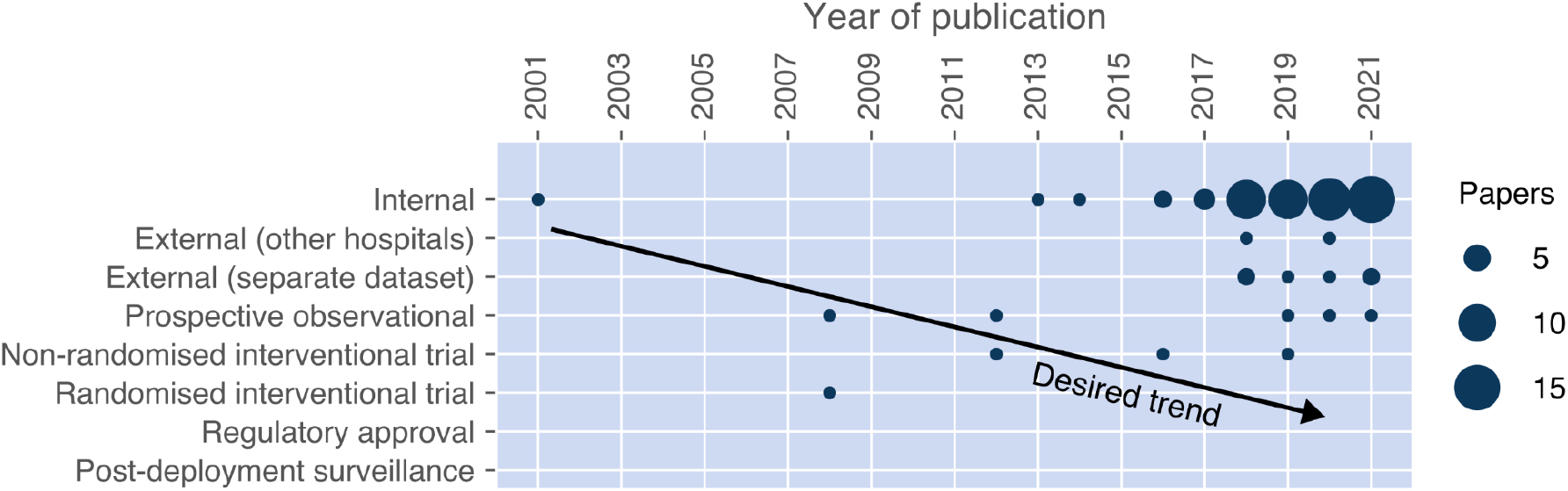
Trends in technology readiness, over time. Where a system was evaluated using multiple methods, the evaluation corresponding to the highest level of technological readiness was counted for each study. The figure format replicates that presented in [9].

A large proportion of the studies (63%) relied on the same database (MIMIC [14]; see Supplementary Figure 3, Additional File 1). The majority of systems used patient demographic data (59, 81%) and measurements collected intermittently over time (68, 93%), with only 7 (10%) systems exploiting physiological waveforms. The number of patient episodes varied greatly between studies using the same datasets (see Supplementary Figure 4, Additional File 1), reflecting their varied inclusion criteria.

### Supervised learning for prediction

Supervised learning is a branch of machine learning which uses labelled datasets consisting of inputs (so-called features, such as patient characteristics) and corresponding labels (e.g. whether that patient develops septic shock in the following 12 hours). Using many pairs of features and labels in a process known as ‘training’, the parameters of a mathematical function are learnt so that the function predicts appropriate outputs given novel inputs (e.g. predicts septic shock risk in a new patient). Common types of function (‘algorithms’) used for this purpose are summarised in Box 1. After training, the learnt function is commonly referred to as a ‘model’.

To evaluate a model’s performance, we ask it to make predictions from unseen (“test”) data, and assess the differences between the predicted and true outcomes. An important goal of the testing phase is to check whether the model has ‘underfitted’ or ‘overfitted’ to the training data: in the former case, the model is too simplistic to capture the true relationship between the features and outcomes. In the latter, the model is too complex relative to the size of the training data, meaning that it simply memorises these data rather than learning patterns that generalise to new patients.

Supervised learning was used in 55 (75%) studies, commonly for the purpose of early identification of septic shock with the aim of enabling timely intervention and reducing disease progression. In particular, 8 studies [15–22] evaluated septic shock prediction systems using the MIMIC dataset, version III. Despite this apparent similarity, however, direct comparison of these systems’ predictive performance is not meaningful due to other sources of heterogeneity: their time between prediction and septic shock onset ranged from 15 minutes to 48 hours, between 25 and 4786 episodes of septic shock were identified during dataset processing, reflecting large differences in inclusion criteria and hence the meaning of the ‘septic shock’ labels, and the studies used different measures of model performance.

#### Box 1.

Common supervised learning algorithms

- <u>Linear regression</u> - a simple approach used to predict a continuous outcome by adding together one or more weighted features. Used in many ‘clinical calculators’ but prone to underfitting, as most relationships in nature are not on a straight line.
- <u>Logistic regression</u> - a non-linear modification of linear regression, often used to predict the probability of a binary outcome.
- <u>K nearest neighbours</u> - prediction of outcome labels based on the most common label of the nearest data points, where K is the number of nearest data points.
- <u>Support vector machines</u> - an approach which uses complicated combinations of features to find a boundary between different outcomes. The learning process is focussed on identifying ‘support vectors’, i.e. data points that are most characteristic of this boundary.
- <u>Decision trees</u> - a rule-based algorithm that separates features into outcomes over successive steps, where each step uses a decision rule learned from the features.
- <u>Random forests</u> - an extension of decision trees in which several trees are built from different samples of the data, and predictions are made using the majority or average vote.
- <u>Artificial neural networks</u> - interconnected layers of artificial ‘neurons’, each of which integrates information using a weighted combination of the activity of earlier neurons in the network. Stacking the layers results in a ‘deep’ neural network that can learn to represent complex information (e.g. images) very efficiently. Training (‘deep learning’) is done by feeding back the error the output neurons made in the outcome prediction and asking successive sets of neurons to adjust their weighted combinations, a process called ‘backpropagation’.

Supervised algorithms were also applied to guide interventions; either indirectly, by predicting urine output response to fluid [23] and mean arterial pressure response to noradrenaline [24], or directly, by guiding weaning [10] or dose regulation of vasopressors [25].

Clinical machine learning systems are often limited by the breadth of data they have access to in comparison to healthcare professionals [26]. Mitra and colleagues [16] found superior prediction of septic shock when using clinical features as compared to features available only from administrative data. Other included studies used supervised algorithms specifically suited to analysing time-series data (recurrent neural networks and hidden Markov models) [17,27–30] and text notes [19], reflecting the way human physicians retain awareness of patient trajectory and clinical context. Four studies sought to use information from physiological waveforms: an earlier study [31] predicted response to fluids based on manually-defined waveform features (pulse pressure variation), whilst later studies [32–34] relied on the algorithm’s ability to identify novel informative features of the waveforms which humans are typically not able to comprehend in real time.

A minority of studies considered integration of supervised learning models into existing clinical workstreams. These included an ‘open-loop’ model in which clinicians’ decisions were fed back into the system to allow them to influence the model predictions [35], software to allow clinicians with no coding experience to create their own predictive tools [36], and a system that calculated the trade-off between cost-effectiveness and prediction accuracy [37].

Several studies compared the performance of multiple algorithms on a single task [38,39], or combined them in “ensemble” models as a means of increasing overall performance and generalisability [15,22,40]. Two studies [41,42] deployed existing models in new hospitals, but first ‘fine tuned’ the models with local data from those hospitals — a technique known as transfer learning. In both cases, transfer learning improved prediction of septic shock, indicating that it can be used to improve model generalisation to new settings.

### Unsupervised learning for phenotype discovery

In contrast to supervised learning, unsupervised algorithms are not provided with labels. Instead, they are designed to discover structure in data. *Clustering* is a popular unsupervised technique which splits data into groups based on common characteristics, such as a shared phenotype of a disease or syndrome [43]. Researchers usually attempt to demonstrate that clusters add new information to a dataset (for example, facilitating imputation of missing clinical data [44,45]), are reproducible in other datasets, or argue for the biological plausibility or prognostic utility of the data clusters.

Several studies used clustering in combination with other machine learning techniques: three [25,46,47] used clustering with supervised learning, aiming to capture phenotypes associated with different responses to cardiovascular treatment. One study [46] aimed to link unsupervised techniques with treatment recommendations more directly: having identified multiple underlying phenotypes in a population of patients with sepsis, the investigators showed that responses to fluid resuscitation differed between phenotypes. It should be emphasised, however, that a causal relationship was not demonstrated here.

### Reinforcement learning for treatment recommendation

Reinforcement learning (RL) is a family of algorithms in which an ‘agent’ (a computer program) learns a strategy for making decisions (a ‘policy’) which achieves a desired outcome. Box 2 describes a hypothetical clinical RL system. RL is able to optimise complex, sequential decision-making, and therefore has the potential to enhance resuscitation [48].

All of the included RL studies focus on a scenario wherein the agent administers intravenous fluids and vasopressors to virtual patients with sepsis, learning the policy (dosing strategy) associated with desirable outcomes. The behaviour of this agent depends on three key elements: the representation of the patient’s clinical status (the “states”), the treatment options available to the agent (the “actions”) and the definition of successful resuscitation (the “reward”).

### Representing patient state

Early studies [49,50] defined patient state using their demographic information, plus their clinical observations and laboratory values from the current time, and input it directly into the RL algorithm. Alternatively, clustering techniques were applied to these data to define discrete patient states [51]. However, these approaches did not take into account trajectories over time and could lead to inconsistent recommendations. Therefore, later studies typically condensed patients’ historical data into a single state which preserved information about their clinical trajectory [52–54].

#### Box 2.

Case study: a hypothetical clinical reinforcement learning system

An RL system is used to select antimicrobial agents for patients with new-onset sepsis, as well as recommending when a trial of cessation would be appropriate. The system is trained on historical patient data (including microbiological samples and sensitivities) with the goal of reducing the requirement for sepsis-related organ support. This system could improve antimicrobial stewardship by optimising for a patient-centred and clinically-important outcome, as opposed to adopting the shorter-term triggers (e.g. recent high temperature) that clinicians frequently use. It also has the potential to identify, and act on, longer-term trends in sensitivities and clinical responses to antibiotics that are only apparent from large volumes of health data.

### Defining treatment goals

Many of the studies defined successful resuscitation using patient-centred outcomes such as survival at 30 or 90 days [47,51,54–57]. However, these outcomes only occur once per patient episode, resulting in a sparse reward signal that makes training difficult and raises concerns over the weak causal link between treatment decisions and an outcome months into the future [58,59]. To overcome this, some studies defined shorter-term rewards such as changes in SOFA score or clinical parameters [53,60,61], but these may not translate into outcomes that are important to patients. More recent studies refined their reward signals further, using them to nudge the agent’s treatment strategy away from rare [62] or unsafe [60,63] decisions.

### Deployment and other challenges

Clinical RL agents are usually trained “offline” (that is, they learn by observing retrospective data rather than experimenting in the real clinical environment). In this setting, it is impossible to know how patients’ outcomes might have changed if they had been treated differently, and model performance must be estimated through statistical methods [64]. Due to the extreme heterogeneity in model architectures, algorithms used, definitions (of states, actions and rewards), and performance measures, it was not possible to directly compare the performance of RL models.

### Learning with biological models

Four studies [11,12,61,65] proposed systems containing biological models augmented with machine learning. These attempted to constrain the way the systems learn from data using explicit mathematical descriptions of physiology or pathophysiology, so that the system recommendations were compatible with our biological knowledge.

Two studies [11,12] linked fluid resuscitation recommendations in sepsis to the output of the FloTrac™ cardiac output monitor. FloTrac™ uses the physiological principle that pulse pressure is proportional to stroke volume via the combined effects of vascular resistance and compliance. It then uses a regression model learnt from multiple patients’ data to predict this relationship for a given patient. Unlike more invasive cardiac output monitors, studies using FloTrac™ were eligible for inclusion in this review as it uses only routinely-collected ICU data (invasive arterial blood pressure and patient demographics).

Two studies [61,65] modelled the cardiovascular system in sepsis as an electrical circuit analogue, wherein charge corresponds to blood volume, current to blood flow, and voltage to blood pressure. Systemic vascular resistance and arterial compliance are represented as a parallel resistor and capacitor, respectively, in a model known as a two-element Windkessel [66]. One of these studies [65] presented a system for titration of noradrenaline infusions in septic patients, which coupled a supervised learning model to a 2-element Windkessel for blood pressure forecasting. The other [61] presented a complex reinforcement learning approach to sepsis cardiovascular resuscitation with IV fluids and vasopressors, combining three neural networks and a two-element Windkessel. In this latter study, however, it is plausible that the neural networks learnt to simply invert the Windkessel model and thereafter learnt in a way that was unconstrained by biological knowledge.

## Conclusions

We performed a scoping review of data-driven systems for personalised cardiovascular resuscitation in sepsis. The systems performed varied tasks, but most commonly used supervised learning to predict septic shock or reinforcement learning to recommend doses of IV fluids and vasopressors. Whilst supervised learning systems can (under favourable conditions) predict events during normal patient care, they rely on humans to operationalise the predictions into treatment decisions that could improve outcomes. The relationship between usual-care events and optimal treatments may be complex: for example, where a future prescription of vasopressors is predicted in a patient with sepsis, the optimal treatment strategy may be earlier control of infection source, which may in turn make future vasopressors unnecessary. Reinforcement learning systems avoid these issues by recommending treatments directly, but offline training using electronic health record data makes them vulnerable to confounding.

The paucity of studies making treatment recommendations solely based on unsupervised clustering may reflect concerns over clinical interpretation of the clusters, which necessarily occurs *post hoc* and is often speculative. However, clusters derived from routinely-collected patient data may be more transparent to most clinicians than those derived from ‘omics’ or cytokine data, as they can inspect familiar variables which most differentiate the clusters.

Traditional biological models are based on strict assumptions and population average parameters. They can therefore show poor predictive performance when applied to individual patients. Augmenting biological models with machine learning may enable more personalised predictions. However, care must be taken to ensure that these augmented systems are still constrained by physiological principles.

There is substantial recent interest in applying data-driven approaches to cardiovascular treatment in sepsis. However, with some notable exceptions, the vast majority of systems to date exist only as proof of concept, with little prospective validation and significant barriers to translation.

## Supporting information

Additional File 2 (extracted study data)

PRISMA-ScR checklist

## Data Availability

All extracted study data are included in Additional file 2.

## List of abbreviations

EMBASE: Excerpta Medica Database
ICU: Intensive care unit
IQR: Interquartile range
MEDLINE: Medical Literature Analysis and Retrieval System Online
MIMIC: Medical Information Mart for Intensive Care
PRESS: Peer review of electronic search strategies
PRISMA-ScR: Preferred reporting items for systematic reviews and meta-analyses, extension for scoping reviews
RL: Reinforcement learning
SOFA: Sequential organ failure assessment

## Declarations

### Ethics approval and consent to participate

Not applicable.

### Consent for publication

Not applicable.

### Availability of data and materials

All extracted study data are included in Additional file 2.

### Authors’ contributions

MK conceived the study. FJRC, MN, PF, ZB and MK designed and executed the search and assessed studies for inclusion. FJRC, PF, ZB and MK extracted data. FJRC and PF performed the initial analysis with all authors contributing to interpretation of data. FJRC, MN, PF, ZB and MK drafted the initial version of the manuscript with all authors contributing to critical revision of the manuscript for important intellectual content and approving the final version. FJRC is the study guarantor.

## Acknowledgements

The authors acknowledge the NIHR Biomedical Research Centre based at Imperial College Healthcare NHS Trust and Imperial College London. The views expressed are those of the authors and not necessarily those of the NHS, the NIHR or the Department of Health.

## References

1 Singer M, Deutschman CS, Seymour CW, et al. The Third International Consensus Definitions for Sepsis and Septic Shock (Sepsis-3). JAMA 2016;315:801–10.

2 Shafique M, Naseer M, Theocharides T, et al. Robust Machine Learning Systems: Challenges, Current Trends, Perspectives, and the Road Ahead. arXiv [cs.CR]. 2021.http://arxiv.org/abs/2101.02559

3 Tricco AC, Lillie E, Zarin W, et al. PRISMA Extension for Scoping Reviews (PRISMA-ScR): Checklist and Explanation. Ann Intern Med 2018;169:467–73.

4 Bone RC, Balk RA, Cerra FB, et al. Definitions for sepsis and organ failure and guidelines for the use of innovative therapies in sepsis. The ACCP/SCCM Consensus Conference Committee. American College of Chest Physicians/Society of Critical Care Medicine. Chest 1992;101:1644–55.

5 Levy MM, Fink MP, Marshall JC, et al. 2001 SCCM/ESICM/ACCP/ATS/SIS International Sepsis Definitions Conference. Intensive Care Med 2003;29:530–8.

6 McGowan J, Sampson M, Salzwedel DM, et al. PRESS Peer Review of Electronic Search Strategies: 2015 Guideline Statement. J Clin Epidemiol 2016;75:40–6.

7 Higgins JPT, Thomas J, Chandler J, et al., editors. Technical Supplement to Chapter 4: Searching for and selecting studies. In: Cochrane Handbook for Systematic Reviews of Interventions Version 6. Cochrane 2021. 62.

8 Kastner M, Wilczynski NL, Walker-Dilks C, et al. Age-specific search strategies for Medline. J Med Internet Res 2006;8:e25.

9 van de Sande D, van Genderen ME, Huiskens J, et al. Moving from bytes to bedside: a systematic review on the use of artificial intelligence in the intensive care unit. Intensive Care Med 2021;47:750–60.

10 Merouani M, Guignard B, Vincent F, et al. Norepinephrine weaning in septic shock patients by closed loop control based on fuzzy logic. Crit Care 2008;12:R155.

11 Khwannimit B, Bhurayanontachai R. Prediction of fluid responsiveness in septic shock patients: comparing stroke volume variation by FloTrac/Vigileo and automated pulse pressure variation. Eur J Anaesthesiol 2012;29:64–9.

12 Krige A, Bland M, Fanshawe T. Fluid responsiveness prediction using Vigileo FloTrac measured cardiac output changes during passive leg raise test. J Intensive Care Med 2016;4:63.

13 Giannini HM, Ginestra JC, Chivers C, et al. A Machine Learning Algorithm to Predict Severe Sepsis and Septic Shock: Development, Implementation, and Impact on Clinical Practice. Crit Care Med 2019;47:1485–92.

14 Johnson AEW, Pollard TJ, Shen L, et al. MIMIC-III, a freely accessible critical care database. Sci Data 2016;3:160035.

15 Darwiche A, Mukherjee S. Machine Learning Methods for Septic Shock Prediction. In: Proceedings of the 2018 International Conference on Artificial Intelligence and Virtual Reality. New York, NY, USA: : Association for Computing Machinery 2018. 104–10.

16 Mitra A, Ashraf K. Sepsis Prediction and Vital Signs Ranking in Intensive Care Unit Patients. arXiv [cs.LG]. 2018.http://arxiv.org/abs/1812.06686

17 Fagerström J, Bång M, Wilhelms D, et al. LiSep LSTM: A Machine Learning Algorithm for Early Detection of Septic Shock. Sci Rep 2019;9:15132.

18 Liu R, Greenstein JL, Granite SJ, et al. Data-driven discovery of a novel sepsis pre-shock state predicts impending septic shock in the ICU. Sci Rep 2019;9:6145.

19 Liu R, Greenstein JL, Sarma SV, et al. Natural Language Processing of Clinical Notes for Improved Early Prediction of Septic Shock in the ICU. Conf Proc IEEE Eng Med Biol Soc 2019;2019:6103–8.

20 Yee CR, Narain NR, Akmaev VR, et al. A Data-Driven Approach to Predicting Septic Shock in the Intensive Care Unit. Biomed Inform Insights 2019;11:1178222619885147.

21 Mollura M, Romano S, Mantoan G, et al. Prediction of Septic Shock Onset in ICU by Instantaneous Monitoring of Vital Signs^*^. In: 2020 42nd Annual International Conference of the IEEE Engineering in Medicine Biology Society (EMBC). 2020. 2768–71.

22 Darwiche A, EL-Geneidy A, Mukherjee S. Improving Septic Shock Prediction with AdaBoost and Cox Regression Model. In: 2021 IEEE International Conference on Consumer Electronics and Computer Engineering (ICCECE). 2021. 522–7.

23 Lin P-C, Huang H-C, Komorowski M, et al. A machine learning approach for predicting urine output after fluid administration. Comput Methods Programs Biomed 2019;177:155–9.

24 Tang Y, Brown S, Sorensen J, et al. Reduced Rank Least Squares for Real-Time Short Term Estimation of Mean Arterial Blood Pressure in Septic Patients Receiving Norepinephrine. IEEE J Transl Eng Health Med 2019;7:4100209.

25 Liu J, Gong M, Guo W, et al. Dose regulation model of norepinephrine based on LSTM network and clustering analysis in sepsis. Proc Int Joint Conf Bioinforma Syst Biol Intell Comput 2020;13:717.

26 Catling FJR, Wolff AH. Temporal convolutional networks allow early prediction of events in critical care. J Am Med Inform Assoc 2020;27:355–65.

27 Ghosh S, Li J, Cao L, et al. Septic shock prediction for ICU patients via coupled HMM walking on sequential contrast patterns. J Biomed Inform 2017;66:19–31.

28 Khoshnevisan F, Ivy J, Capan M, et al. Recent Temporal Pattern Mining for Septic Shock Early Prediction. In: 2018 IEEE International Conference on Healthcare Informatics (ICHI). 2018. 229–40.

29 Lin C, Zhang Y, Ivy J, et al. Early Diagnosis and Prediction of Sepsis Shock by Combining Static and Dynamic Information Using Convolutional-LSTM. In: 2018 IEEE International Conference on Healthcare Informatics (ICHI). 2018. 219–28.

30 Khoshnevisan F, Chi M. An Adversarial Domain Separation Framework for Septic Shock Early Prediction Across EHR Systems. In: 2020 IEEE International Conference on Big Data (Big Data). 2020. 64–73.

31 Oliveira-Costa CDA de, Friedman G, Vieira SRR, et al. Pulse pressure variation and prediction of fluid responsiveness in patients ventilated with low tidal volumes. Clinics 2012;67:773–8.

32 Brown SM, Sorensen J, Lanspa MJ, et al. Multi-complexity measures of heart rate variability and the effect of vasopressor titration: a prospective cohort study of patients with septic shock. BMC Infect Dis 2016;16:551.

33 Kamaleswaran R, Lian J, Lin D-L, et al. Predicting Volume Responsiveness Among Sepsis Patients Using Clinical Data and Continuous Physiological Waveforms. AMIA Annu Symp Proc 2020;2020:619–28.

34 Mollura M, Lehman L-WH, Mark RG, et al. A novel artificial intelligence based intensive care unit monitoring system: using physiological waveforms to identify sepsis. Philosophical Transactions of the Royal Society A: Mathematical, Physical and Engineering Sciences 2021;379:20200252.

35 Gupta A, Liu T, Crick C. Utilizing time series data embedded in electronic health records to develop continuous mortality risk prediction models using hidden Markov models: A sepsis case study. Stat Methods Med Res 2020;29:3409–23.

36 La Cava W, Williams H, Fu W, et al. Evaluating recommender systems for AI-driven biomedical informatics. Bioinformatics 2021;37:250–6.

37 Robinson DP, Saria S. Trading-off cost of deployment versus accuracy in learning predictive models. In: Proceedings of the Twenty-Fifth International Joint Conference on Artificial Intelligence. AAAI Press 2016. 1974–82.

38 Misra D, Avula V, Wolk DM, et al. Early Detection of Septic Shock Onset Using Interpretable Machine Learners. J Clin Med Res 2021;10. doi:10.3390/jcm10020301

39 Chen Y-M, Kao Y, Hsu C-C, et al. Real-time interactive artificial intelligence of things-based prediction for adverse outcomes in adult patients with pneumonia in the emergency department. Acad Emerg Med 2021;28:1277–85.

40 Kim J, Chang H, Kim D, et al. Machine learning for prediction of septic shock at initial triage in emergency department. J Crit Care 2020;55:163–70.

41 Mao Q, Jay M, Hoffman JL, et al. Multicentre validation of a sepsis prediction algorithm using only vital sign data in the emergency department, general ward and ICU. BMJ Open 2018;8:e017833.

42 Wardi G, Carlile M, Holder A, et al. Predicting Progression to Septic Shock in the Emergency Department Using an Externally Generalizable Machine-Learning Algorithm. Ann Emerg Med 2021;77:395–406.

43 Seymour CW, Kennedy JN, Wang S, et al. Derivation, Validation, and Potential Treatment Implications of Novel Clinical Phenotypes for Sepsis. JAMA 2019;321:2003–17.

44 Ho JC, Lee CH, Ghosh J. Septic Shock Prediction for Patients with Missing Data. ACM Trans Manage Inf Syst 2014;5:1–15.

45 Yang X, Zhang Y, Chi M. Time-aware Subgroup Matrix Decomposition: Imputing Missing Data Using Forecasting Events. In: 2018 IEEE International Conference on Big Data (Big Data). 2018. 1524–33.

46 Zhang Z, Zhang G, Goyal H, et al. Identification of subclasses of sepsis that showed different clinical outcomes and responses to amount of fluid resuscitation: a latent profile analysis. Crit Care 2018;22:347.

47 Ma P, Liu J, Shen F, et al. Individualized resuscitation strategy for septic shock formalized by finite mixture modeling and dynamic treatment regimen. Crit Care 2021;25:243.

48 Liu S, See KC, Ngiam KY, et al. Reinforcement Learning for Clinical Decision Support in Critical Care: Comprehensive Review. J Med Internet Res 2020;22:e18477.

49 Raghu A, Komorowski M, Ahmed I, et al. Deep Reinforcement Learning for Sepsis Treatment. arXiv e-prints 2017;:arXiv:1711.09602.

50 Raghu A, Komorowski M, Celi LA, et al. Continuous State-Space Models for Optimal Sepsis Treatment -a Deep Reinforcement Learning Approach. arXiv [cs.LG]. 2017.http://arxiv.org/abs/1705.08422

51 Komorowski M, Celi LA, Badawi O, et al. The Artificial Intelligence Clinician learns optimal treatment strategies for sepsis in intensive care. Nat Med 2018;24:1716–20.

52 Peng X, Ding Y, Wihl D, et al. Improving Sepsis Treatment Strategies by Combining Deep and Kernel-Based Reinforcement Learning. AMIA Annu Symp Proc 2018;2018:887–96.

53 Jeter R, Lehman L-W, Josef C, et al. Learning to treat hypotensive episodes in sepsis patients using a counterfactual reasoning framework. bioRxiv. 2021. doi:10.1101/2021.03.03.21252863

54 Zhu S, Pu J. A self-supervised method for treatment recommendation in sepsis. Frontiers of Information Technology & Electronic Engineering 2021;22:926–39.

55 Killian TW, Zhang H, Subramanian J, et al. An Empirical Study of Representation Learning for Reinforcement Learning in Healthcare. arXiv [cs.LG]. 2020.http://arxiv.org/abs/2011.11235

56 Zhang Z, Zheng B, Liu N. Individualized fluid administration for critically ill patients with sepsis with an interpretable dynamic treatment regimen model. Sci Rep 2020;10:1–9.

57 Roggeveen L, El Hassouni A, Ahrendt J, et al. Transatlantic transferability of a new reinforcement learning model for optimizing haemodynamic treatment for critically ill patients with sepsis. Artif Intell Med 2021;112:102003.

58 Yu C, Ren G, Liu J. Deep Inverse Reinforcement Learning for Sepsis Treatment. In: 2019 IEEE International Conference on Healthcare Informatics (ICHI). 2019. 1–3.

59 Lu M, Shahn Z, Sow D, et al. Is Deep Reinforcement Learning Ready for Practical Applications in Healthcare? A Sensitivity Analysis of Duel-DDQN for Hemodynamic Management in Sepsis Patients. arXiv [cs.LG]. 2020.http://arxiv.org/abs/2005.04301

60 Jia Y, Lawton T, Burden J, et al. Safety-driven design of machine learning for sepsis treatment. J Biomed Inform 2021;117:103762.

61 Nanayakkara T, Clermont G, Langmead CJ, et al. Unifying cardiovascular modelling with deep reinforcement learning for uncertainty aware control of sepsis treatment. PLOS Digital Health 2022;1:e0000012.

62 Satija H, Thomas PS, Pineau J, et al. Multi-Objective SPIBB: Seldonian Offline Policy Improvement with Safety Constraints in Finite MDPs. arXiv [cs.LG]. 2021.http://arxiv.org/abs/2106.00099

63 Festor P, Luise G, Komorowski M, et al. Enabling risk-aware Reinforcement Learning for medical interventions through uncertainty decomposition. arXiv [cs.AI]. 2021.http://arxiv.org/abs/2109.07827

64 Levine S, Kumar A, Tucker G, et al. Offline Reinforcement Learning: Tutorial, Review, and Perspectives on Open Problems. arXiv [cs.LG]. 2020.http://arxiv.org/abs/2005.01643

65 Tang Y, Brown SM, Sorensen J, et al. Physiology-Informed Real-Time Mean Arterial Blood Pressure Learning and Prediction for Septic Patients Receiving Norepinephrine. IEEE Trans Biomed Eng 2021;68:181–91.

66 Shi Y, Lawford P, Hose R. Review of zero-D and 1-D models of blood flow in the cardiovascular system. Biomed Eng Online 2011;10:33.

