## Additional File 1 (appendices and supplementary figures) for "Data-driven decision support for individualised cardiovascular resuscitation in sepsis: a scoping review and primer for clinicians"

**Appendix A: Literature search strategy (Ovid MEDLINE)** 2
**Appendix B: Data collection form** 5
**Supplementary figures** 7

### Appendix A: Literature search strategy (Ovid MEDLINE)

1. sepsis/ or shock, septic/ or endotoxemia/
2. sepsis.ti,ab.
3. septic*.ti,ab.
4. 1 or 2 or 3
5. exp Artificial Intelligence/
6. Latent Class Analysis/
7. exp neural networks, computer/
8. Models, Cardiovascular/
9. Patient-Specific Modeling/
10. decision making, computer-assisted/ or exp diagnosis, computer-assisted/ or therapy, computer-assisted/ or drug therapy, computer-assisted/
11. Decision Support Systems, Clinical/
12. mediation analysis/
13. markov chains/
14. causal inference.ti,ab.
15. dynamic treatment.ti,ab.
16. controller.ti,ab.
17. closed loop.ti,ab.
18. digital twin.ti,ab.
19. machine learning.ti,ab.
20. artificial intelligence.ti,ab.
21. supervised.ti,ab.
22. unsupervised.ti,ab.
23. reinforcement.ti,ab.
24. big data.ti,ab.
25. data driven.ti,ab.
26. markov.ti,ab.
27. pomdp.ti,ab.
28. mdp.ti,ab.
29. hmm.ti,ab.
30. (classifi* or classify*).ti.
31. (regress or regression or regressor*).ti,ab.
32. deep learn*.ti,ab.
33. neural net*.ti,ab.
34. ((recurrent or convolutional or feedforward) adj net*).ti,ab.
35. k means.ti,ab.
36. principle component*.ti,ab.
37. nearest neighbour?.ti,ab.
38. support vector?.ti,ab.

39. random forest?.ti,ab.
40. gradient boost*.ti,ab.
41. bayes*.ti,ab.
42. ((comput* or mathematic* or cardiac* or vascular* or cardio* or h?emodynamic* or dynamic* or physiolog* or mechanis* or biolog*) adj2 (model or models or model?ing or model?ed)).ti,ab.
43. (cluster? or clustering).ti,ab.
44. fuzzy logic.ti,ab.
45. 5 or 6 or 7 or 8 or 9 or 10 or 11 or 12 or 13 or 14 or 15 or 16 or 17 or 18 or 19 or 20 or 21 or 22 or 23 or 24 or 25 or 26 or 27 or 28 or 29 or 30 or 31 or 32 or 33 or 34 or 35 or 36 or 37 or 38 or 39 or 40 or 41 or 42 or 43 or 44
46. Fluid Therapy/
47. Vasoconstrictor Agents/
48. Cardiotonic Agents/
49. Drug Therapy, Computer-Assisted/
50. crystalloid?.ti,ab.
51. ((IV or intravenous* or parenteral* or resuscitat* or giving) adj1 fluid?).ti,ab.
52. (fluid? adj2 (prescri* or therap* or treatment? or resuscitat* or medication? or administ*)).ti,ab.
53. colloid?.ti,ab.
54. vasopressor?.ti,ab.
55. (vasoactives or (vasoactive adj (prescri* or treatment* or therap* or drug? or medication?))).ti,ab.
56. (noradrenaline or norepinephrine).ti,ab.
57. dopamine.ti,ab.
58. vasopressin.ti,ab.
59. phenylephrine.ti,ab.
60. metaraminol.ti,ab.
61. inotrop*.ti,ab.
62. (adrenaline or epinephrine).ti,ab.
63. dobutamine.ti,ab.
64. milrinone.ti,ab.
65. levosimendan.ti,ab.
66. ((personali* or individuali* or "patient specific" or guide* or determine* or recommend* or assist*) adj2 (prescri* or therap* or treatment? or medic* or resuscitat*)).ti,ab.
67. 46 or 47 or 48 or 49 or 50 or 51 or 52 or 53 or 54 or 55 or 56 or 57 or 58 or 59 or 60 or 61 or 62 or 63 or 64 or 65 or 66
68. 4 and 45 and 67
69. (predict* adj2 "septic shock").ti,ab.
70. (diagnos* adj2 "septic shock").ti,ab.
71. (detect* adj2 "septic shock").ti,ab.
72. (anticipat* adj2 "septic shock").ti,ab.
73. (recogni* adj2 "septic shock").ti,ab.
74. 69 or 70 or 71 or 72 or 73

75. 74 and 45
76. 68 or 75
77. (exp animal/ or nonhuman/) not exp human/
78. 76 not 77
79. exp adult/
80. adult.mp. or middle aged.sh. or age:.tw.
81. 79 or 80
82. (child: or adolescent or infan:).mp.
83. (infan: or child:).mp. or gestation:.tw.
84. 82 or 83
85. 84 not 81
86. 78 not 85

### Appendix B: Data collection form

General information

- Title (string)
- Year of publication (integer)
- Funding source (categorical, i.e. choose one)
  - Academic
  - Commercial
  - Mixed
  - None
  - Unclear or not reported

System type

- System type (Boolean for each of the below, i.e. can choose multiple)
  - Supervised learning
  - Unsupervised learning
  - Reinforcement learning
  - Biological / physiological models
  - Algorithm used
- Algorithms used (string)

Study data

- Datasets used (Boolean for each of the below, i.e. can choose multiple)
  - MIMIC (any version)
  - eICU Collaborative Research Database
  - Private dataset / datasets
  - Other public dataset
  - Name/names of other public dataset/datasets
- Number of hospitals is specified (Boolean)
- Total number of hospitals (integer)
- Number of patient episodes is specified (Boolean)
- Total number of patient episodes (integer)
- Data resolution (Boolean for each of the below, i.e. can choose multiple)
  - Non-temporal
  - Temporal, non-waveform
  - Temporal, waveform

Definitions

- Sepsis (Boolean for each of the below, i.e. can choose multiple)
  - Sepsis definition is specified
  - Defined using Sepsis-3

    - Defined using 'Infection + systemic inflammatory response syndrome'
    - Defined using clinical codes
    - Defined using other criteria
- Septic shock (Boolean for each of the below, i.e. can choose multiple)
    - Septic shock definition is specified
    - Definition requires vasopressors or inotropes
    - Definition requires initial fluid resuscitation
    - Definition requires elevated lactate

Validation

- Extent of validation (Boolean for each of the below, i.e. can choose multiple)
    - Internal validation
    - External retrospective validation (other hospitals)
    - External retrospective validation (separate dataset)
    - Prospective observational validation
    - Non-randomised interventional validation (clinical trial)
    - Randomised interventional validation (clinical trial)
    - Regulatory approval
    - Post-deployment surveillance

Other

- Any other comments (string)

### Supplementary figures

**Supplementary Figure 1: Study funding sources over time.**

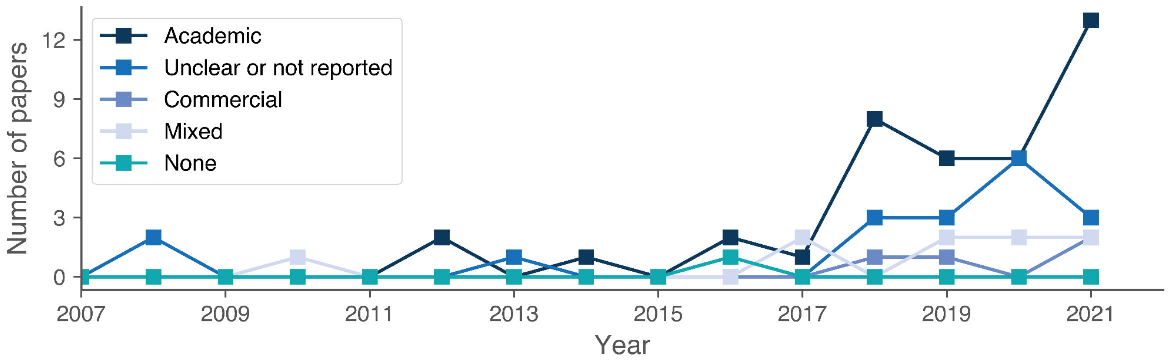


**Supplementary Figure 2: Sepsis and septic shock definitions used.** The cells on the diagonal show the proportion of all papers using the corresponding definition. The other cells show the proportion of papers using that combination of definitions.

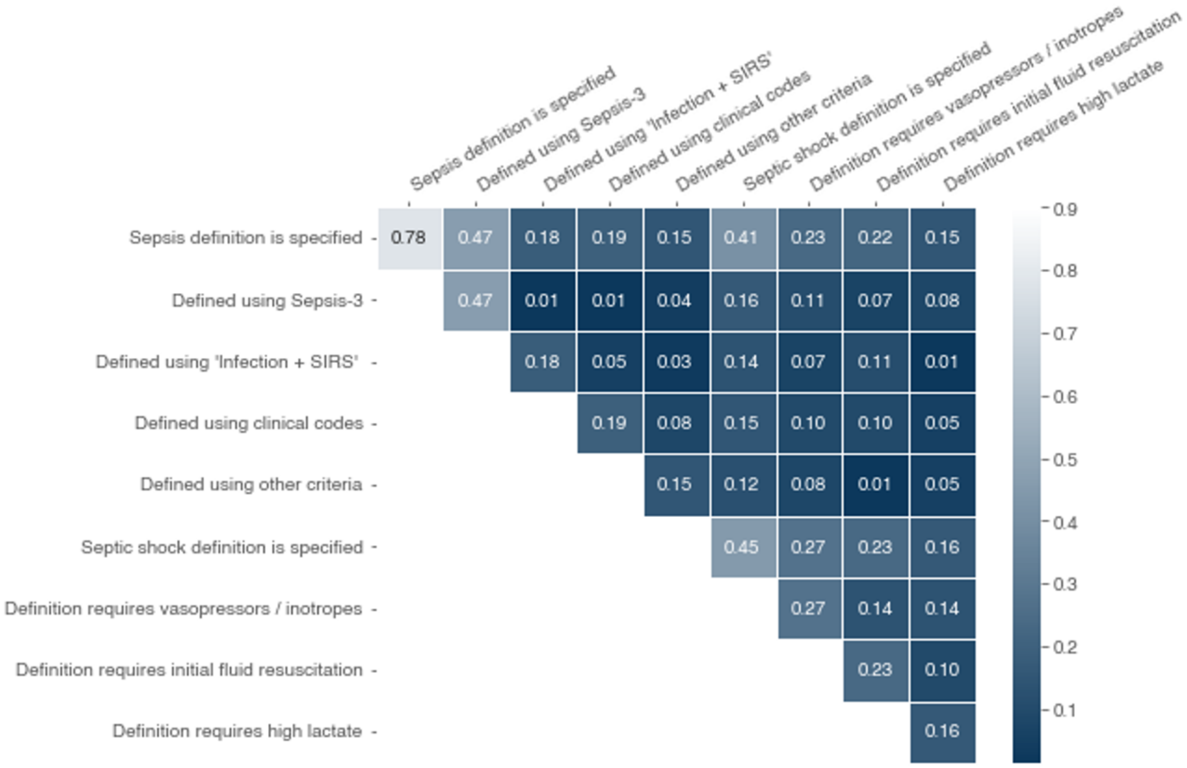

**Supplementary Figure 3: Types of dataset used.** Note that, as some studies use more than one dataset, the percentages sum to more than 100%.

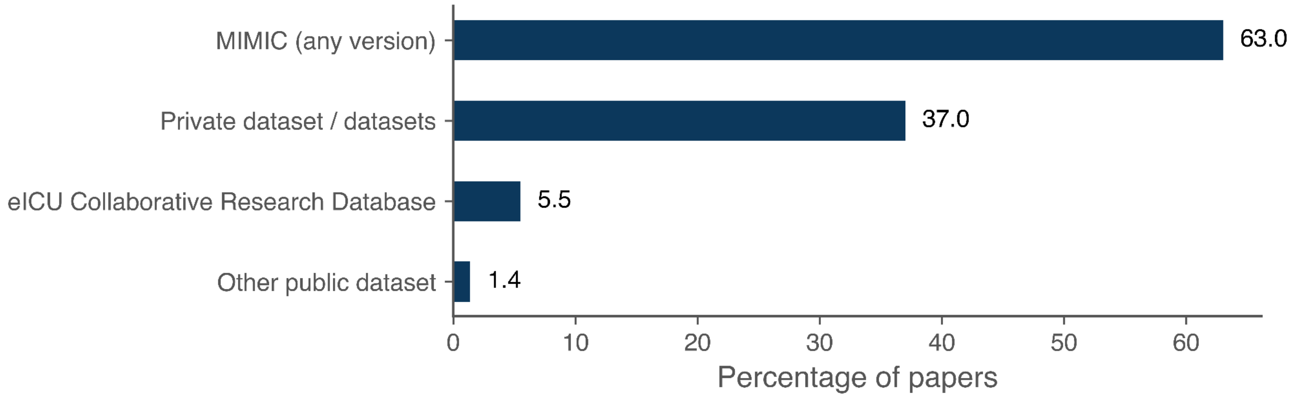


**Supplementary Figure 4: Number patient episodes, stratified by dataset used.**

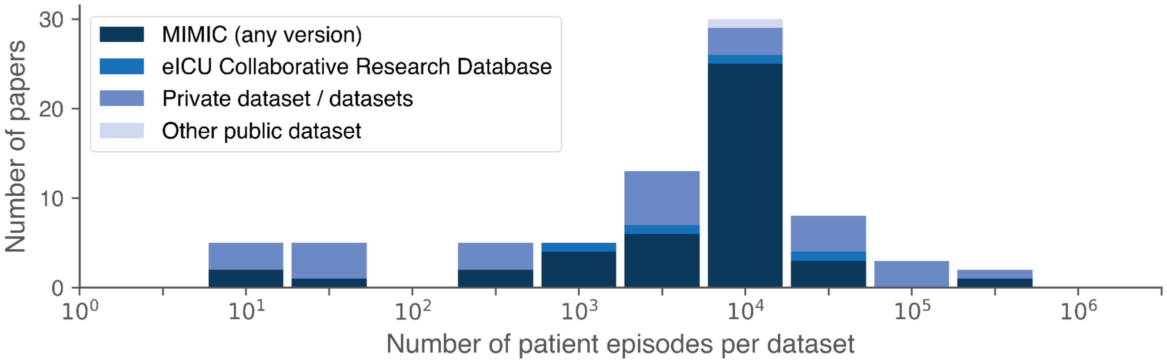
